## Supplemental Material for "Pasteurized retail dairy enables genomic surveillance of H5N1 avian influenza virus in United States cattle"

**S1 Fig: Nextclade clustering of HA segment against the H5Nx 2.3.4.4 clade tree.**

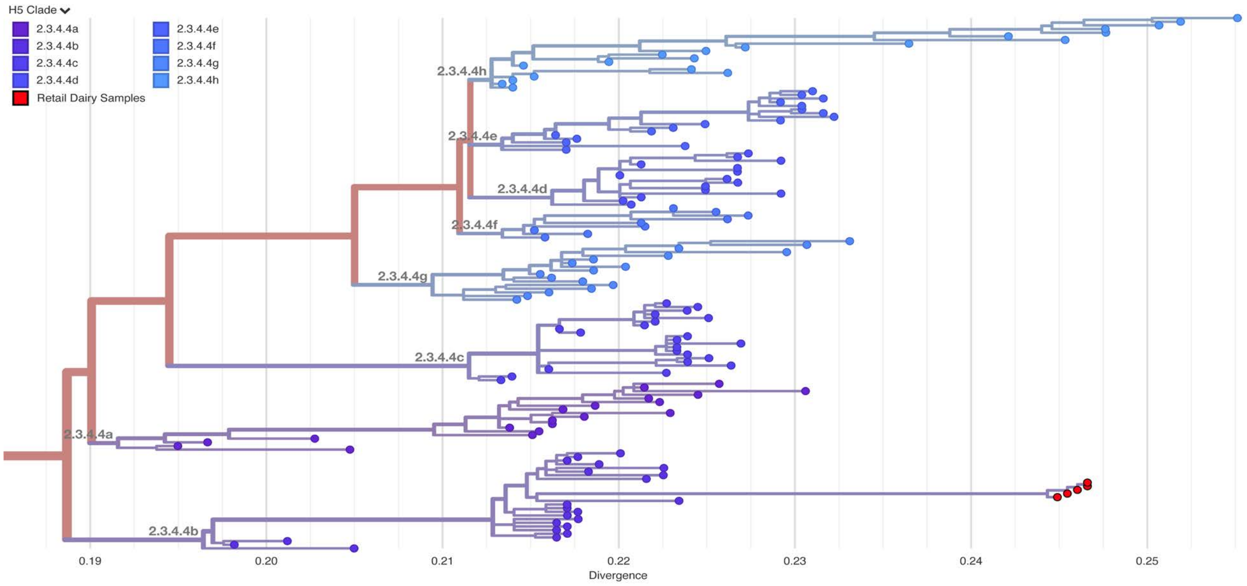


**Supplemental data 1:**

To test primer specificity *in silico*, several H5 HA sequences from birds [OR858836.1 (goose, NY 2023), OR819057.1 (peregrine falcon, NY 2022), OQ968076.1 (goose, WY 2022), OQ968052.1 (goose, MN 2022), OQ968012.1 (chicken, WI 2022)] were downloaded and the two sets of selected primers and probes were aligned to ensure they would bind to other 2.3.4.4b sequences from the current outbreak in North American birds. This was done to confirm the primers would continue working in the event of a new introduction from birds to cows. Sequences from A/Victoria/4897/2022 H1N1 and A/Darwin/9/2021 H3N2, two isolates that have been recommended as vaccine components in recent years, were also downloaded to ensure the selected primers and probes would not align to commonly-circulating sequences (47). The primer sequences did not align to sequences from influenza B viruses B/Austria/13594/7/2021 (B/Victoria lineage) and B/Phuket/3073/2013 (B/Yamagata lineage). To test this specificity in vitro, the two primer/probe sets were tested on H5N1 clade 2.3.4.4b vRNA, A/California/04/2009(H1N1) vRNA, and Influenza A/Kawasaki/173/2001 M gene RNA transcript. Only one set proved specific to the H5 sequence, with the primer sequence 5'-GGGAAGCTATGCGACCTAAAT-3' (forward) and 5'-CATTCCGGCACTCTGATGAA-3' (reverse) and the probe sequence 5'-ACATTGGGTTTCCGAGGAGCCATC-3' with a FAM fluorophore. This set was used for all further analysis.

**S1 Table: Sequences from Andersen Lab github used for generating tiled-amplicon scheme.**

| Sequence |
|---|
| Consensus_SRR28752446_HA_cns_threshold_0.5_quality_20 |
| Consensus_SRR28752447_HA_cns_threshold_0.5_quality_20 |

| |
|---|
| Consensus_SRR28752448_HA_cns_threshold_0.5_quality_20 |
| Consensus_SRR28752449_HA_cns_threshold_0.5_quality_20 |
| Consensus_SRR28752450_HA_cns_threshold_0.5_quality_20 |
| Consensus_SRR28752451_HA_cns_threshold_0.5_quality_20 |
| Consensus_SRR28752452_HA_cns_threshold_0.5_quality_20 |
| Consensus_SRR28752453_HA_cns_threshold_0.5_quality_20 |
| Consensus_SRR28752454_HA_cns_threshold_0.5_quality_20 |
| Consensus_SRR28752455_HA_cns_threshold_0.5_quality_20 |
| Consensus_SRR28752456_HA_cns_threshold_0.5_quality_20 |
| Consensus_SRR28752457_HA_cns_threshold_0.5_quality_20 |
| Consensus_SRR28752458_HA_cns_threshold_0.5_quality_20 |
| Consensus_SRR28752459_HA_cns_threshold_0.5_quality_20 |
| Consensus_SRR28752460_HA_cns_threshold_0.5_quality_20 |
| Consensus_SRR28752461_HA_cns_threshold_0.5_quality_20 |
| Consensus_SRR28752462_HA_cns_threshold_0.5_quality_20 |
| Consensus_SRR28752463_HA_cns_threshold_0.5_quality_20 |
| Consensus_SRR28752464_HA_cns_threshold_0.5_quality_20 |
| Consensus_SRR28752465_HA_cns_threshold_0.5_quality_20 |
| Consensus_SRR28752466_HA_cns_threshold_0.5_quality_20 |
| Consensus_SRR28752467_HA_cns_threshold_0.5_quality_20 |
| Consensus_SRR28752468_HA_cns_threshold_0.5_quality_20 |
| Consensus_SRR28752469_HA_cns_threshold_0.5_quality_20 |
| Consensus_SRR28752470_HA_cns_threshold_0.5_quality_20 |
| Consensus_SRR28752471_HA_cns_threshold_0.5_quality_20 |
| Consensus_SRR28752472_HA_cns_threshold_0.5_quality_20 |
| Consensus_SRR28752473_HA_cns_threshold_0.5_quality_20 |
| Consensus_SRR28752474_HA_cns_threshold_0.5_quality_20 |
| Consensus_SRR28752475_HA_cns_threshold_0.5_quality_20 |
| Consensus_SRR28752476_HA_cns_threshold_0.5_quality_20 |
| Consensus_SRR28752477_HA_cns_threshold_0.5_quality_20 |
| Consensus_SRR28752478_HA_cns_threshold_0.5_quality_20 |
| Consensus_SRR28752479_HA_cns_threshold_0.5_quality_20 |
| Consensus_SRR28752480_HA_cns_threshold_0.5_quality_20 |
| Consensus_SRR28752481_HA_cns_threshold_0.5_quality_20 |
| Consensus_SRR28752482_HA_cns_threshold_0.5_quality_20 |
| Consensus_SRR28752483_HA_cns_threshold_0.5_quality_20 |
| Consensus_SRR28752484_HA_cns_threshold_0.5_quality_20 |
| Consensus_SRR28752485_HA_cns_threshold_0.5_quality_20 |
| Consensus_SRR28752486_HA_cns_threshold_0.5_quality_20 |
| Consensus_SRR28752487_HA_cns_threshold_0.5_quality_20 |
| Consensus_SRR28752488_HA_cns_threshold_0.5_quality_20 |
| Consensus_SRR28752489_HA_cns_threshold_0.5_quality_20 |
| Consensus_SRR28752490_HA_cns_threshold_0.5_quality_20 |
| Consensus_SRR28752491_HA_cns_threshold_0.5_quality_20 |
| Consensus_SRR28752492_HA_cns_threshold_0.5_quality_20 |

| |
|---|
| Consensus_SRR28752493_HA_cns_threshold_0.5_quality_20 |
| Consensus_SRR28752494_HA_cns_threshold_0.5_quality_20 |
| Consensus_SRR28752495_HA_cns_threshold_0.5_quality_20 |
| Consensus_SRR28752496_HA_cns_threshold_0.5_quality_20 |
| Consensus_SRR28752497_HA_cns_threshold_0.5_quality_20 |
| Consensus_SRR28752498_HA_cns_threshold_0.5_quality_20 |
| Consensus_SRR28752499_HA_cns_threshold_0.5_quality_20 |
| Consensus_SRR28752500_HA_cns_threshold_0.5_quality_20 |
| Consensus_SRR28752501_HA_cns_threshold_0.5_quality_20 |
| Consensus_SRR28752502_HA_cns_threshold_0.5_quality_20 |
| Consensus_SRR28752503_HA_cns_threshold_0.5_quality_20 |
| Consensus_SRR28752504_HA_cns_threshold_0.5_quality_20 |
| Consensus_SRR28752505_HA_cns_threshold_0.5_quality_20 |
| Consensus_SRR28752506_HA_cns_threshold_0.5_quality_20 |
| Consensus_SRR28752507_HA_cns_threshold_0.5_quality_20 |
| Consensus_SRR28752508_HA_cns_threshold_0.5_quality_20 |
| Consensus_SRR28752509_HA_cns_threshold_0.5_quality_20 |
| Consensus_SRR28752510_HA_cns_threshold_0.5_quality_20 |
| Consensus_SRR28752511_HA_cns_threshold_0.5_quality_20 |
| Consensus_SRR28752512_HA_cns_threshold_0.5_quality_20 |
| Consensus_SRR28752513_HA_cns_threshold_0.5_quality_20 |
| Consensus_SRR28752514_HA_cns_threshold_0.5_quality_20 |
| Consensus_SRR28752515_HA_cns_threshold_0.5_quality_20 |
| Consensus_SRR28752516_HA_cns_threshold_0.5_quality_20 |
| Consensus_SRR28752517_HA_cns_threshold_0.5_quality_20 |
| Consensus_SRR28752518_HA_cns_threshold_0.5_quality_20 |
| Consensus_SRR28752519_HA_cns_threshold_0.5_quality_20 |
| Consensus_SRR28752520_HA_cns_threshold_0.5_quality_20 |
| Consensus_SRR28752521_HA_cns_threshold_0.5_quality_20 |
| Consensus_SRR28752522_HA_cns_threshold_0.5_quality_20 |
| Consensus_SRR28752523_HA_cns_threshold_0.5_quality_20 |
| Consensus_SRR28752524_HA_cns_threshold_0.5_quality_20 |
| Consensus_SRR28752525_HA_cns_threshold_0.5_quality_20 |
| Consensus_SRR28752526_HA_cns_threshold_0.5_quality_20 |
| Consensus_SRR28752527_HA_cns_threshold_0.5_quality_20 |
| Consensus_SRR28752528_HA_cns_threshold_0.5_quality_20 |
| Consensus_SRR28752529_HA_cns_threshold_0.5_quality_20 |
| Consensus_SRR28752530_HA_cns_threshold_0.5_quality_20 |
| Consensus_SRR28752531_HA_cns_threshold_0.5_quality_20 |
| Consensus_SRR28752532_HA_cns_threshold_0.5_quality_20 |
| Consensus_SRR28752533_HA_cns_threshold_0.5_quality_20 |
| Consensus_SRR28752534_HA_cns_threshold_0.5_quality_20 |
| Consensus_SRR28752535_HA_cns_threshold_0.5_quality_20 |
| Consensus_SRR28752536_HA_cns_threshold_0.5_quality_20 |
| Consensus_SRR28752537_HA_cns_threshold_0.5_quality_20 |

| |
|---|
| Consensus_SRR28752538_HA_cns_threshold_0.5_quality_20 |
| Consensus_SRR28752539_HA_cns_threshold_0.5_quality_20 |
| Consensus_SRR28752540_HA_cns_threshold_0.5_quality_20 |
| Consensus_SRR28752541_HA_cns_threshold_0.5_quality_20 |
| Consensus_SRR28752542_HA_cns_threshold_0.5_quality_20 |
| Consensus_SRR28752543_HA_cns_threshold_0.5_quality_20 |
| Consensus_SRR28752544_HA_cns_threshold_0.5_quality_20 |
| Consensus_SRR28752545_HA_cns_threshold_0.5_quality_20 |
| Consensus_SRR28752546_HA_cns_threshold_0.5_quality_20 |
| Consensus_SRR28752547_HA_cns_threshold_0.5_quality_20 |
| Consensus_SRR28752548_HA_cns_threshold_0.5_quality_20 |
| Consensus_SRR28752549_HA_cns_threshold_0.5_quality_20 |
| Consensus_SRR28752550_HA_cns_threshold_0.5_quality_20 |
| Consensus_SRR28752551_HA_cns_threshold_0.5_quality_20 |
| Consensus_SRR28752552_HA_cns_threshold_0.5_quality_20 |
| Consensus_SRR28752553_HA_cns_threshold_0.5_quality_20 |
| Consensus_SRR28752554_HA_cns_threshold_0.5_quality_20 |
| Consensus_SRR28752555_HA_cns_threshold_0.5_quality_20 |
| Consensus_SRR28752556_HA_cns_threshold_0.5_quality_20 |
| Consensus_SRR28752557_HA_cns_threshold_0.5_quality_20 |
| Consensus_SRR28752558_HA_cns_threshold_0.5_quality_20 |
| Consensus_SRR28752559_HA_cns_threshold_0.5_quality_20 |
| Consensus_SRR28752560_HA_cns_threshold_0.5_quality_20 |
| Consensus_SRR28752561_HA_cns_threshold_0.5_quality_20 |
| Consensus_SRR28752562_HA_cns_threshold_0.5_quality_20 |
| Consensus_SRR28752563_HA_cns_threshold_0.5_quality_20 |
| Consensus_SRR28752564_HA_cns_threshold_0.5_quality_20 |
| Consensus_SRR28752565_HA_cns_threshold_0.5_quality_20 |
| Consensus_SRR28752566_HA_cns_threshold_0.5_quality_20 |
| Consensus_SRR28752567_HA_cns_threshold_0.5_quality_20 |
| Consensus_SRR28752568_HA_cns_threshold_0.5_quality_20 |
| Consensus_SRR28752569_HA_cns_threshold_0.5_quality_20 |
| Consensus_SRR28752570_HA_cns_threshold_0.5_quality_20 |
| Consensus_SRR28752571_HA_cns_threshold_0.5_quality_20 |
| Consensus_SRR28752572_HA_cns_threshold_0.5_quality_20 |
| Consensus_SRR28752573_HA_cns_threshold_0.5_quality_20 |
| Consensus_SRR28752574_HA_cns_threshold_0.5_quality_20 |
| Consensus_SRR28752575_HA_cns_threshold_0.5_quality_20 |
| Consensus_SRR28752576_HA_cns_threshold_0.5_quality_20 |
| Consensus_SRR28752577_HA_cns_threshold_0.5_quality_20 |
| Consensus_SRR28752578_HA_cns_threshold_0.5_quality_20 |
| Consensus_SRR28752579_HA_cns_threshold_0.5_quality_20 |
| Consensus_SRR28752580_HA_cns_threshold_0.5_quality_20 |
| Consensus_SRR28752581_HA_cns_threshold_0.5_quality_20 |
| Consensus_SRR28752582_HA_cns_threshold_0.5_quality_20 |

| |
|---|
| Consensus_SRR28752583_HA_cns_threshold_0.5_quality_20 |
| Consensus_SRR28752584_HA_cns_threshold_0.5_quality_20 |
| Consensus_SRR28752585_HA_cns_threshold_0.5_quality_20 |
| Consensus_SRR28752586_HA_cns_threshold_0.5_quality_20 |
| Consensus_SRR28752587_HA_cns_threshold_0.5_quality_20 |
| Consensus_SRR28752588_HA_cns_threshold_0.5_quality_20 |
| Consensus_SRR28752589_HA_cns_threshold_0.5_quality_20 |
| Consensus_SRR28752590_HA_cns_threshold_0.5_quality_20 |
| Consensus_SRR28752591_HA_cns_threshold_0.5_quality_20 |
| Consensus_SRR28752592_HA_cns_threshold_0.5_quality_20 |
| Consensus_SRR28752593_HA_cns_threshold_0.5_quality_20 |
| Consensus_SRR28752594_HA_cns_threshold_0.5_quality_20 |
| Consensus_SRR28752595_HA_cns_threshold_0.5_quality_20 |
| Consensus_SRR28752596_HA_cns_threshold_0.5_quality_20 |
| Consensus_SRR28752597_HA_cns_threshold_0.5_quality_20 |
| Consensus_SRR28752598_HA_cns_threshold_0.5_quality_20 |
| Consensus_SRR28752599_HA_cns_threshold_0.5_quality_20 |
| Consensus_SRR28752600_HA_cns_threshold_0.5_quality_20 |
| Consensus_SRR28752601_HA_cns_threshold_0.5_quality_20 |
| Consensus_SRR28752602_HA_cns_threshold_0.5_quality_20 |
| Consensus_SRR28752603_HA_cns_threshold_0.5_quality_20 |
| Consensus_SRR28752604_HA_cns_threshold_0.5_quality_20 |
| Consensus_SRR28752605_HA_cns_threshold_0.5_quality_20 |
| Consensus_SRR28752606_HA_cns_threshold_0.5_quality_20 |
| Consensus_SRR28752607_HA_cns_threshold_0.5_quality_20 |
| Consensus_SRR28752608_HA_cns_threshold_0.5_quality_20 |
| Consensus_SRR28752609_HA_cns_threshold_0.5_quality_20 |
| Consensus_SRR28752610_HA_cns_threshold_0.5_quality_20 |
| Consensus_SRR28752611_HA_cns_threshold_0.5_quality_20 |
| Consensus_SRR28752612_HA_cns_threshold_0.5_quality_20 |
| Consensus_SRR28752613_HA_cns_threshold_0.5_quality_20 |
| Consensus_SRR28752614_HA_cns_threshold_0.5_quality_20 |
| Consensus_SRR28752615_HA_cns_threshold_0.5_quality_20 |
| Consensus_SRR28752616_HA_cns_threshold_0.5_quality_20 |
| Consensus_SRR28752617_HA_cns_threshold_0.5_quality_20 |
| Consensus_SRR28752618_HA_cns_threshold_0.5_quality_20 |
| Consensus_SRR28752619_HA_cns_threshold_0.5_quality_20 |
| Consensus_SRR28752620_HA_cns_threshold_0.5_quality_20 |
| Consensus_SRR28752621_HA_cns_threshold_0.5_quality_20 |
| Consensus_SRR28752622_HA_cns_threshold_0.5_quality_20 |
| Consensus_SRR28752623_HA_cns_threshold_0.5_quality_20 |
| Consensus_SRR28752624_HA_cns_threshold_0.5_quality_20 |
| Consensus_SRR28752625_HA_cns_threshold_0.5_quality_20 |
| Consensus_SRR28752626_HA_cns_threshold_0.5_quality_20 |
| Consensus_SRR28752627_HA_cns_threshold_0.5_quality_20 |

| |
|---|
| Consensus_SRR28752628_HA_cns_threshold_0.5_quality_20 |
| Consensus_SRR28752629_HA_cns_threshold_0.5_quality_20 |
| Consensus_SRR28752630_HA_cns_threshold_0.5_quality_20 |
| Consensus_SRR28752631_HA_cns_threshold_0.5_quality_20 |
| Consensus_SRR28752632_HA_cns_threshold_0.5_quality_20 |
| Consensus_SRR28752633_HA_cns_threshold_0.5_quality_20 |
| Consensus_SRR28752634_HA_cns_threshold_0.5_quality_20 |
| Consensus_SRR28752635_HA_cns_threshold_0.5_quality_20 |
| Consensus_SRR28752636_HA_cns_threshold_0.5_quality_20 |
| Consensus_SRR28752637_HA_cns_threshold_0.5_quality_20 |
| Consensus_SRR28752638_HA_cns_threshold_0.5_quality_20 |
| Consensus_SRR28752639_HA_cns_threshold_0.5_quality_20 |
| Consensus_SRR28752640_HA_cns_threshold_0.5_quality_20 |
| Consensus_SRR28752641_HA_cns_threshold_0.5_quality_20 |
| Consensus_SRR28752642_HA_cns_threshold_0.5_quality_20 |
| Consensus_SRR28752643_HA_cns_threshold_0.5_quality_20 |
| Consensus_SRR28752644_HA_cns_threshold_0.5_quality_20 |
| Consensus_SRR28752645_HA_cns_threshold_0.5_quality_20 |
| Consensus_SRR28752646_HA_cns_threshold_0.5_quality_20 |
| Consensus_SRR28752647_HA_cns_threshold_0.5_quality_20 |
| Consensus_SRR28752648_HA_cns_threshold_0.5_quality_20 |
| Consensus_SRR28752649_HA_cns_threshold_0.5_quality_20 |
| Consensus_SRR28752650_HA_cns_threshold_0.5_quality_20 |
| Consensus_SRR28752651_HA_cns_threshold_0.5_quality_20 |
| Consensus_SRR28752652_HA_cns_threshold_0.5_quality_20 |
| Consensus_SRR28752653_HA_cns_threshold_0.5_quality_20 |
| Consensus_SRR28752654_HA_cns_threshold_0.5_quality_20 |
| Consensus_SRR28752655_HA_cns_threshold_0.5_quality_20 |
| Consensus_SRR28752656_HA_cns_threshold_0.5_quality_20 |
| Consensus_SRR28752657_HA_cns_threshold_0.5_quality_20 |
| Consensus_SRR28752658_HA_cns_threshold_0.5_quality_20 |
| Consensus_SRR28752659_HA_cns_threshold_0.5_quality_20 |
| Consensus_SRR28752660_HA_cns_threshold_0.5_quality_20 |
| Consensus_SRR28752661_HA_cns_threshold_0.5_quality_20 |
| Consensus_SRR28752662_HA_cns_threshold_0.5_quality_20 |
| Consensus_SRR28752663_HA_cns_threshold_0.5_quality_20 |
| Consensus_SRR28752664_HA_cns_threshold_0.5_quality_20 |
| Consensus_SRR28752665_HA_cns_threshold_0.5_quality_20 |
| Consensus_SRR28752666_HA_cns_threshold_0.5_quality_20 |
| Consensus_SRR28752667_HA_cns_threshold_0.5_quality_20 |
| Consensus_SRR28752668_HA_cns_threshold_0.5_quality_20 |
| Consensus_SRR28752669_HA_cns_threshold_0.5_quality_20 |
| Consensus_SRR28752670_HA_cns_threshold_0.5_quality_20 |
| Consensus_SRR28752671_HA_cns_threshold_0.5_quality_20 |
| Consensus_SRR28752672_HA_cns_threshold_0.5_quality_20 |

| |
|---|
| Consensus_SRR28752673_HA_cns_threshold_0.5_quality_20 |
| Consensus_SRR28752674_HA_cns_threshold_0.5_quality_20 |
| Consensus_SRR28752675_HA_cns_threshold_0.5_quality_20 |
| Consensus_SRR28752676_HA_cns_threshold_0.5_quality_20 |
| Consensus_SRR28752677_HA_cns_threshold_0.5_quality_20 |
| Consensus_SRR28752678_HA_cns_threshold_0.5_quality_20 |
| Consensus_SRR28752679_HA_cns_threshold_0.5_quality_20 |
| Consensus_SRR28752680_HA_cns_threshold_0.5_quality_20 |
| Consensus_SRR28752681_HA_cns_threshold_0.5_quality_20 |
| Consensus_SRR28752682_HA_cns_threshold_0.5_quality_20 |
| Consensus_SRR28752683_HA_cns_threshold_0.5_quality_20 |
| Consensus_SRR28752684_HA_cns_threshold_0.5_quality_20 |

**S2 Table: Primer pools used in tiled-amplicon sequencing.**

| Name | Pool name | Sequence |
|---|---|---|
| HPAI-HA-250_1_LEFT | 1 | TGGAGAACATAGTACTACTTCTTGCAA |
| HPAI-HA-250_1_RIGHT | 1 | GATGAATTCGTCGCACATTGGG |
| HPAI-HA-250_2_LEFT | 2 | ATGGGGTGAAGCCACTGATTTT |
| HPAI-HA-250_2_RIGHT | 2 | CCCTAGTGATGTTTCATGATTTGGC |
| HPAI-HA-250_3_LEFT | 1 | CACATGTTGAGCAGAATAAATCATTTTGAG |
| HPAI-HA-250_3_RIGHT | 1 | GTCTGCTCTTCTGCATTGTTGG |
| HPAI-HA-250_4_LEFT | 2 | GATGCATACCCAACAATAAAGATAAGCT |
| HPAI-HA-250_4_RIGHT | 2 | TGGATTGCATCATCTGGTTTTAAGATTG |
| HPAI-HA-250_5_LEFT | 1 | GCTACTAGATCCCAAGTAAACGGG |
| HPAI-HA-250_5_RIGHT | 1 | GAGAGGATGTATGTTGTGAAATGGC |
| HPAI-HA-250_6_LEFT | 2 | AAAGTGGAGTGGAATATGGCCA |
| HPAI-HA-250_6_RIGHT | 2 | ACCAACCATCAACCATTCCCTG |
| HPAI-HA-250_7_LEFT | 1 | GAAAAAGAGGTCTGTTTGGGGC |
| HPAI-HA-250_7_RIGHT | 1 | TGTTCAAATTCTCTATCCTCCTTTCTAAGT |
| HPAI-HA-250_8_LEFT | 2 | ACTCAATCATTGACAAAATGAACACTCA |
| HPAI-HA-250_8_RIGHT | 2 | TTCGAAACAGCCGTTACCCAG |
| HPAI-HA-250_9_LEFT | 1 | GTCAAGAACCTTTACGACAAAGTCAG |
| HPAI-HA-250_9_RIGHT | 1 | GGGAACTTGCCGCTGTTGAAT |
| HPAI-MP-250_1_LEFT | 1 | CTAACCGAGGTCGAAACGTACG |
| HPAI-MP-250_1_RIGHT | 1 | CCACTTAGGGCACTTTGGACAA |
| HPAI-MP-250_2_LEFT | 2 | TTGGGATTTGTGTTCACGCTCA |
| HPAI-MP-250_2_RIGHT | 2 | CAATGCCACTTCTGCGGTCA |
| HPAI-MP-250_3_LEFT | 1 | CGGTGCACTTGCCAGTTGTAT |
| HPAI-MP-250_3_RIGHT | 1 | TCCATGGCTTCCACTGCTTG |
| HPAI-MP-250_4_LEFT | 2 | TGGTTTTGGCCAGTACTACAGC |
| HPAI-MP-250_4_RIGHT | 2 | AGATCCCAATGATACTTGCGGC |
| HPAI-MP-250_5_LEFT | 1 | CCTTGAAAATTTGCAGGCCTACC |
| HPAI-MP-250_5_RIGHT | 1 | TCAACATCCACAGCACTCTGC |
| HPAI-NA-250_1_LEFT | 1 | GAATCCAAATCAGAAGATAACAACCATTG |
| HPAI-NA-250_1_RIGHT | 1 | GCCCGCTAATGTTACCGAAGTA |
| HPAI-NA-250_2_LEFT | 2 | GGTAAATCAGACGTATATCAACATCAGC |

| | | |
|---|---|---|
| HPAI-NA-250_2_RIGHT | 2 | GAGTGTTTGTCATTCAGCAGAGC |
| HPAI-NA-250_3_LEFT | 1 | TTGTTATAAGAGAACCATTCATCTCATGC |
| HPAI-NA-250_3_RIGHT | 1 | TCTGGACCAGAAATACCGATTGTC |
| HPAI-NA-250_4_LEFT | 2 | TTTGAGTCTGTTGCTTGGTCGG |
| HPAI-NA-250_4_RIGHT | 2 | TGAAGATTTTATATGAGGCCTGCCC |
| HPAI-NA-250_5_LEFT | 1 | CTCAAGAATCTGAATGTGCTTGCG |
| HPAI-NA-250_5_RIGHT | 1 | ACTCCAGATTTTGATTAAAAGATACCCAC |
| HPAI-NA-250_6_LEFT | 2 | CGGGTGATATTATGTGTGTGTGC |
| HPAI-NA-250_6_RIGHT | 2 | CGCTTCTGGAACTAGTGCTTTTTG |
| HPAI-NA-250_7_LEFT | 1 | GGCATATGGGGTAAAAGGGTTTTC |
| HPAI-NA-250_7_RIGHT | 1 | TAGCTCAACCCAGAAACAAGGC |
| HPAI-NA-250_8_LEFT | 2 | GGACAGTAGTTTCTCAGTGAAGCA |
| HPAI-NA-250_8_RIGHT | 2 | CAATGGTGAATGGCAACTCAGC |
| HPAI-NP-250_1_LEFT | 1 | CTCAAGGCACCAAACGATCCTA |
| HPAI-NP-250_1_RIGHT | 1 | AGCACTGGGATGTTCCTCCA |
| HPAI-NP-250_2_LEFT | 2 | CCAAAACAGCATAACCATAGAAAGGA |
| HPAI-NP-250_2_RIGHT | 2 | GGAATGCCAAATCATCAAGTGAGTG |
| HPAI-NP-250_3_LEFT | 1 | GAGATCAGAAGAATTTGGCGTCAAG |
| HPAI-NP-250_3_RIGHT | 1 | TCCTCGTTTGATCATTCGAATCAATTC |
| HPAI-NP-250_4_LEFT | 2 | TCACTGATGCAAGGCTCAACC |
| HPAI-NP-250_4_RIGHT | 2 | CATTCCCAGGATTCCGGCTTT |
| HPAI-NP-250_5_LEFT | 1 | TCAAGGGAAAGCTCCAAACAGC |
| HPAI-NP-250_5_RIGHT | 1 | TGGAGCAGACGGAAAGGATCA |
| HPAI-NP-250_6_LEFT | 2 | ACTTGCTGTAGCCAGTGGATATG |
| HPAI-NP-250_6_RIGHT | 2 | GCAATCTGAACTCCTCTGGTGG |
| HPAI-NP-250_7_LEFT | 1 | GAGAGTGTCAAGCTTCATCAGAGG |
| HPAI-NP-250_7_RIGHT | 1 | TGATGGTTGCTCTCTCGAATGG |
| HPAI-NP-250_8_LEFT | 2 | TCTGCAGGACAAATCAGCGTAC |
| HPAI-NP-250_8_RIGHT | 2 | CATTGTTCATGTCAAAGGAAGGCA |
| HPAI-NS-250_1_LEFT | 1 | CCAACACTGTGTTAAGCTTTCAGG |
| HPAI-NS-250_1_RIGHT | 1 | GGCACAGAGGCAATAGTCATTTT |
| HPAI-NS-250_2_LEFT | 2 | TGCTGGGAAGCAGATAGTGGA |
| HPAI-NS-250_2_RIGHT | 2 | TCCAGCCGATTGAAGATCACAC |
| HPAI-NS-250_3_LEFT | 1 | TCCCTCAGTATCAGAATGGACCA |
| HPAI-NS-250_3_RIGHT | 1 | AAAGTTTCAGAGACTCGAACTGTGT |
| HPAI-NS-250_4_LEFT | 2 | TGCCTTCTCTTCCAGGACATACT |
| HPAI-NS-250_4_RIGHT | 2 | TCTGTGATCTTTAGTCTGTGTCGC |
| HPAI-PA-250_1_LEFT | 1 | GGAAGACTTTGTGCGACAATGC |
| HPAI-PA-250_1_RIGHT | 1 | GCTCGGTCTCTCCCTTCGATTA |
| HPAI-PA-250_2_LEFT | 2 | GTCTGTTTCATGTATTCGGATTTCCA |
| HPAI-PA-250_2_RIGHT | 2 | TGAACTTCCCTTCGCGTTACTC |
| HPAI-PA-250_3_LEFT | 1 | CCTCCCTGATTTGTATGACTACAGAG |
| HPAI-PA-250_3_RIGHT | 1 | AGGAATCCCATAGACCTCTACTGG |
| HPAI-PA-250_4_LEFT | 2 | TGATGAAGAGAGCAGAGCAAGAA |
| HPAI-PA-250_4_RIGHT | 2 | AGAAAGCTTGCCCTCAATGCA |

| | | |
|---|---|---|
| HPAI-PA-250_5_LEFT | 1 | ACCGAACTTCTCCAGCCTTGA |
| HPAI-PA-250_5_RIGHT | 1 | TATGCCCTCCCCCTCATGACTA |
| HPAI-PA-250_6_LEFT | 2 | ATTACCTGATGGGCCTCCCT |
| HPAI-PA-250_6_RIGHT | 2 | GGAATTTTCTCCTCATTTTCAATGTCTTG |
| HPAI-PA-250_7_LEFT | 1 | TGAGAAAGGCATAAACCCTAATTACCT |
| HPAI-PA-250_7_RIGHT | 1 | TTCACTCTGAATCCAGCTTGCT |
| HPAI-PA-250_8_LEFT | 2 | AGATGTTAGCGATCTAAGACAGTACG |
| HPAI-PA-250_8_RIGHT | 2 | GGGCTGTGTTTATGTATACTCCCTT |
| HPAI-PA-250_9_LEFT | 1 | GGAACTATTTCACAGCGGAGGT |
| HPAI-PA-250_9_RIGHT | 1 | GTTAGAGAGAATTCCATGCTCACAAAG |
| HPAI-PA-250_10_LEFT | 2 | GGAGGACAAATCTGTATGGATTCATT |
| HPAI-PA-250_10_RIGHT | 2 | CTCATCTCCATGCCCCATTTCA |
| HPAI-PA-250_11_LEFT | 1 | TGCTCCTACGGACTGCAATAGG |
| HPAI-PA-250_11_RIGHT | 1 | TTCCCAATAGAGCCTTCCTCCA |
| HPAI-PA-250_12_LEFT | 2 | GGCCGAATCTTCTGTCAAAGAGA |
| HPAI-PA-250_12_RIGHT | 2 | CCAAGATCGAAGGTTCCAGGTT |
| HPAI-PB1-250_1_LEFT | 1 | CCTTACTCTTCTTGAAAGTTCCAGC |
| HPAI-PB1-250_1_RIGHT | 1 | CGCAGTCCGTTTGTGCATAC |
| HPAI-PB1-250_2_LEFT | 2 | GGCACCTCAACTCAATCCAATTG |
| HPAI-PB1-250_2_RIGHT | 2 | CCTCTATAGTATTAGCTAATGCAGTTGC |
| HPAI-PB1-250_3_LEFT | 1 | GAGTGGACAAGTTGACCCAAGG |
| HPAI-PB1-250_3_RIGHT | 1 | CCGTTGTGTGACCATTTTCTTGG |
| HPAI-PB1-250_4_LEFT | 2 | GTGGTGGAATCAATGGATAAAGAGGA |
| HPAI-PB1-250_4_RIGHT | 2 | AGTACACAAACCCTCTGATTTGCA |
| HPAI-PB1-250_5_LEFT | 1 | TGACAAAAGACGCCGAAAGAGG |
| HPAI-PB1-250_5_RIGHT | 1 | CTGGTTCTCATTCCATTTAGTGTTGTC |
| HPAI-PB1-250_6_LEFT | 2 | GTGAGAAAGATGATGACTAATTCGCAA |
| HPAI-PB1-250_6_RIGHT | 2 | CTGCCGGTATTTGTGTTCGAAG |
| HPAI-PB1-250_7a_LEFT | 1 | CAGGAATGTATTGAGCATTGCACC |
| HPAI-PB1-250_7a_RIGHT | 1 | CCCAGAACTGTACTCAGCATGTT |
| HPAI-PB1-250_7b_LEFT | 1 | GGAATGTATTGAGCATTGCACC |
| HPAI-PB1-250_7b_RIGHT | 1 | CCCAGAACTGTACTCAGCATG |
| HPAI-PB1-250_8_LEFT | 2 | CCTCTTCTAATAGATGGTACGGCC |
| HPAI-PB1-250_8_RIGHT | 2 | GCTCATATTGATTCCTACCAGCTTG |
| HPAI-PB1-250_9_LEFT | 1 | GTGAATGCTCCAAATCATGAGGG |
| HPAI-PB1-250_9_RIGHT | 1 | TTGTTGATCATGTTGTTCTTTATCACTGT |
| HPAI-PB1-250_10_LEFT | 2 | GAGTTGCCCAGCTTTGGAGTT |
| HPAI-PB1-250_10_RIGHT | 2 | TCCTGGTTTTGAGCGGGTTTG |
| HPAI-PB1-250_11_LEFT | 1 | GGAGACACACAAATTCAAACAAGGAG |
| HPAI-PB1-250_11_RIGHT | 1 | GCATCACCACAGCATTGTTCAC |
| HPAI-PB1-250_12_LEFT | 2 | GGGAAGGCTTTGTAATCCCCTG |
| HPAI-PB1-250_12_RIGHT | 2 | CTCCTGTATGAACTGCTAGGGAAG |
| HPAI-PB1-250_13_LEFT | 1 | CGCTCTATTCTTAATACAAGCCAAAGG |
| HPAI-PB1-250_13_RIGHT | 1 | ATTTCTGCCGTCTGAGCTCTTC |
| HPAI-PB2-250_1_LEFT | 1 | TGGAGAGAATAAAAGAACTGAGAGATCT |

| | | |
|---|---|---|
| HPAI-PB2-250_1_RIGHT | 1 | GATCCGGCATCATTTGTCTTGC |
| HPAI-PB2-250_2_LEFT | 2 | TATCCAATCGCAGCAGACAAGC |
| HPAI-PB2-250_2_RIGHT | 2 | TGGTTTCTGAAGTGTACAGGGC |
| HPAI-PB2-250_3_LEFT | 1 | ACAATTCACTATCCAAAGGTATACAAAACC |
| HPAI-PB2-250_3_RIGHT | 1 | CCTGGAGTTCTTCTTTCTTTTCCTTTG |
| HPAI-PB2-250_4_LEFT | 2 | TTCCAAATGAAGTGGGAGCGAG |
| HPAI-PB2-250_4_RIGHT | 2 | CGTTTCTCACTTCTCCTCCTGG |
| HPAI-PB2-250_5_LEFT | 1 | GTGTCTACATTGAGGTGCTGCA |
| HPAI-PB2-250_5_RIGHT | 1 | CATATGTCCACGGCTTGTTCCT |
| HPAI-PB2-250_6_LEFT | 2 | CCACAGCACACAAATTGGAGGA |
| HPAI-PB2-250_6_RIGHT | 2 | CGTTGCTCTTCTTCCAACCATAGT |
| HPAI-PB2-250_7_LEFT | 1 | GTCAAAAGGGAAGAAGAGGTGCT |
| HPAI-PB2-250_7_RIGHT | 1 | GTTGACAAAGTTCAGATCACCTCG |
| HPAI-PB2-250_8_LEFT | 2 | GAAGCAATAATCGTGGCCATGG |
| HPAI-PB2-250_8_RIGHT | 2 | TGACTCTTATTCCCCTCAGTGACA |
| HPAI-PB2-250_9_LEFT | 1 | TGACAATGTGATGGGAATGATCGG |
| HPAI-PB2-250_9_RIGHT | 1 | TGATCTCCCACATCATTGATGACG |
| HPAI-PB2-250_10_LEFT | 2 | TCACCCGAAGAAGTCAGCGA |
| HPAI-PB2-250_10_RIGHT | 2 | TGTCCTCACGAACCCACTGTAT |
| HPAI-PB2-250_11_LEFT | 1 | AAGATGGAGTTCGAGCCATTCC |
| HPAI-PB2-250_11_RIGHT | 1 | TTGAACACTGGAGAATTGCCTCTT |
| HPAI-PB2-250_12_LEFT | 2 | TGCAATTCTCCTCTCTGACTGTG |
| HPAI-PB2-250_12_RIGHT | 2 | TGCTCAGCTCATTGATGCTCAG |
| HPAI-PB2-250_13_LEFT | 1 | CGAAGATCCAGATGAAGGCACA |
| HPAI-PB2-250_13_RIGHT | 1 | TGGCCATCCGAATTCTTTTGGT |
| H5_HPAI-PB2-250-0-LEFT | 2 | ATATACGCGTAGCRAAAGCAGGTCAA |
| H5_HPAI-PB2-250-0-RIGHT | 2 | CTTGCTCCAGAGGGTTTGTCC |
| H5_HPAI-PB2-250-14-LEFT | 2 | TGAAGGCACAGCTGGAGTGG |
| H5_HPAI-PB2-250-14-RIGHT | 2 | GCGTAGTAGAAACAAGGTCG |
| H5_HPAI-PB1-250-0-LEFT | 2 | ATATACGCGTAGCRAAAGCAGGCAAA |
| H5_HPAI-PB1-250-0-RIGHT | 2 | CCACTTGGCTCATTGTCATCAGG |
| H5_HPAI-PB1-250-14-LEFT | 2 | ACGAACAGATGTATCAAAAGTGCTGC |
| H5_HPAI-PB1-250-14-RIGHT | 2 | ATATACGCGTAGTAGAAACAAGGCAT |
| H5_HPAI-PA-250-0-LEFT | 2 | ACGCGTAGCAAAAGCAGGTACTGAT |
| H5_HPAI-PA-250-0-RIGHT | 2 | ACTATTCACCACTGTCCAGGCC |
| H5_HPAI-PA-250-13-LEFT | 1 | ATATGCATCTCCTCAACTCGAGGG |
| H5_HPAI-PA-250-13-RIGHT | 1 | ACGCGTAGTAGAAACAAGGTACTTTTTTGGACAG |
| H5_HPAI-HA-250-0-LEFT | 2 | ATATACGCGTAGCRAAAGCAGGGGTTCACTCTG |
| H5_HPAI-HA-250-0-RIGHT | 2 | TTTCCGAGGAGCCATCCAGC |
| H5_HPAI-HA-250-10-LEFT | 2 | GGAAAGTGTGARAAATGGGACG |
| H5_HPAI-HA-250-10-RIGHT | 2 | ATATACGCGTAGTAGAAACAAGGGTG |
| H5_HPAI-NP-250-0-LEFT | 2 | ATATACGCGTAGCAAAAGCAGGGTAGATAATCAC |
| H5_HPAI-NP-250-0-RIGHT | 2 | CTCATCAAATGCCGAGAGAACC |
| H5_HPAI-NP-250-9-LEFT | 1 | CAACAGAGAGCATCTGCAGGAC |
| H5_HPAI-NP-250-9-RIGHT | 1 | ATATACGCGTAGTAGAAACAAGGGTATT |

| | | |
|---|---|---|
| H5_HPAI-NA-250-0-LEFT | 2 | ATATACGCGTAGCRAAAGCAGGAGTTC |
| H5_HPAI-NA-250-0-RIGHT | 2 | GCCTGCTCAGCAAGAAAATTGG |
| H5_HPAI-NA-250-9-LEFT | 1 | ACTGGTCAGGATATAGTGGGAG |
| H5_HPAI-NA-250-9-RIGHT | 1 | ATATACGCGTAGTAGAAACAAGGAGT |
| H5_HPAI-MP-250-0-LEFT | 2 | ATATACGCGTAGCRAAAGCAGGTAGATATTG |
| H5_HPAI-MP-250-0-RIGHT | 2 | AGCGTCTACGCTGCAGTCC |
| H5_HPAI-MP-250-6-LEFT | 2 | AAACGGATGGGAGTGCAAC |
| H5_HPAI-MP-250-6-RIGHT | 2 | ATATACGCGTAGTAGAAACAAGGTAG |
| H5_HPAI-NS-250-0-LEFT | 2 | ATATACGCGTAGCRAAAGCAGGGTGACA |
| H5_HPAI-NS-250-0-RIGHT | 2 | TCGTCGGATTCTTCCTCCAGAATC |
| H5_HPAI-NS-250-5-LEFT | 1 | TACAGAGATTCGCTTGGAGAAGC |
| H5_HPAI-NS-250-5-RIGHT | 1 | ATATACGCGTAGTAGAAACAAGGGTGTT |

**S3 Table: Full results from RT-qPCR testing.**

| carton | date purchased | date expiration | average cycle threshold | primer set | processing plant state |
|---|---|---|---|---|---|
| dholab_carton_0001 | 4/24/24 | 5/8/24 | | M gene | IA |
| dholab_carton_0002 | 4/24/24 | 5/1/24 | | M gene | IA |
| dholab_carton_0003 | 4/24/24 | 6/5/24 | | M gene | MN |
| dholab_carton_0004 | 4/24/24 | 5/30/24 | | M gene | MN |
| dholab_carton_0005 | 4/24/24 | 6/20/24 | 32.435 | M gene | CO |
| dholab_carton_0006 | 4/24/24 | 5/19/24 | 33.055 | M gene | CO |
| dholab_carton_0007 | 4/24/24 | 5/8/24 | | M gene | WI |
| dholab_carton_0008 | 4/24/24 | 5/8/24 | | M gene | WI |
| dholab_carton_0009 | 4/24/24 | 5/2/24 | | M gene | WI |
| dholab_carton_0010 | 4/24/24 | 5/1/24 | | M gene | IA |
| dholab_carton_0011 | 4/24/24 | 5/1/24 | | M gene | IA |
| dholab_carton_0012 | 4/24/24 | 5/2/24 | | M gene | WI |
| dholab_carton_0013 | 4/24/24 | 5/7/24 | | M gene | WI |
| dholab_carton_0014 | 4/24/24 | | | M gene | IA |
| dholab_carton_0015 | 4/26/24 | | | M gene | WI |
| dholab_carton_0016 | 5/2/24 | 5/17/24 | | M gene | VA |
| dholab_carton_0017 | 5/2/24 | 6/6/24 | | M gene | NY |
| dholab_carton_0018 | 5/2/24 | 6/4/24 | | M gene | OH |
| dholab_carton_0019 | 5/2/24 | 6/17/24 | 33.52 | M gene | KY |
| dholab_carton_0020 | 5/2/24 | 6/27/24 | | M gene | CO |
| dholab_carton_0021 | 5/2/24 | 6/4/24 | 31.17 | M gene | TX |
| dholab_carton_0022 | 5/2/24 | 5/16/24 | | M gene | UT |
| dholab_carton_0023 | 5/2/24 | 5/30/24 | | M gene | IN |
| dholab_carton_0024 | 5/2/24 | 7/11/24 | 26.29 | M gene | MI |
| dholab_carton_0005 | 4/24/24 | 6/20/24 | 31.34 | M gene | CO |
| dholab_carton_0020 | 5/2/24 | 6/27/24 | | H5-specific | CO |
| dholab_carton_0005 | 4/24/24 | 6/20/24 | 32.205 | H5-specific | CO |
| dholab_carton_0024 | 5/2/24 | 7/11/24 | 27.365 | H5-specific | MI |

| | | | | | |
|---|---|---|---|---|---|
| dholab_carton_0025 | 5/20/24 | 7/29/24 | | H5-specific | ID |
| dholab_carton_0026 | 5/20/24 | 7/2/24 | | H5-specific | KY |
| dholab_carton_0027 | 5/20/24 | 7/27/24 | 38.435 | H5-specific | IN |
| dholab_carton_0028 | 5/20/24 | 7/12/24 | 68.48 | H5-specific | KS |
| dholab_carton_0029 | 5/20/24 | 6/21/24 | | H5-specific | FL |
| dholab_carton_0030 | 5/20/24 | 8/12/24 | | H5-specific | MI |
| dholab_carton_0031 | 5/20/24 | 11/18/24 | | H5-specific | MO |
| dholab_carton_0032 | 5/20/24 | 6/5/24 | | H5-specific | WI |
| dholab_carton_0033 | 5/20/24 | 9/12/24 | 38.785 | H5-specific | CO |
| dholab_carton_0034 | 5/20/24 | 6/24/24 | | H5-specific | NC |
| dholab_carton_0035 | 5/20/24 | 7/4/24 | | H5-specific | CA |
| dholab_carton_0036 | 5/20/24 | 7/8/24 | 36.79 | H5-specific | CO |
| dholab_carton_0037 | 5/20/24 | 6/30/24 | | H5-specific | NY |
| dholab_carton_0038 | 5/20/24 | 6/20/24 | | H5-specific | OR |
| dholab_carton_0039 | 5/20/24 | 6/11/24 | | H5-specific | OH |
| dholab_carton_0041 | 5/10/24 | | 35.905 | H5-specific | MI |
| dholab_carton_0042 | 5/10/24 | | | H5-specific | MI |
| dholab_carton_0024 | 5/2/24 | 7/11/24 | 28.03 | H5-specific | MI |
| dholab_carton_0043 | 6/5/24 | 6/23/24 | | H5-specific | MN |
| dholab_carton_0044 | 6/5/24 | 6/13/24 | | H5-specific | WI |
| dholab_carton_0045 | 6/5/24 | 6/11/24 | | H5-specific | IN |
| dholab_carton_0046 | 6/5/24 | 8/16/24 | 31.71 | H5-specific | CO |
| dholab_carton_0047 | 6/5/24 | 6/6/24 | | H5-specific | MI |
| dholab_carton_0048 | 6/5/24 | 8/22/24 | 33.535 | H5-specific | MI |
| dholab_carton_0049 | 6/5/24 | 6/13/24 | | H5-specific | IA |
| dholab_carton_0050 | 6/5/24 | 6/1/24 | | H5-specific | IL |
| dholab_carton_0051 | 6/5/24 | 6/10/24 | | H5-specific | IL |
| dholab_carton_0052 | 6/5/24 | 6/18/24 | | H5-specific | WI |
| dholab_carton_0053 | 6/5/24 | 6/28/24 | | H5-specific | IN |
| dholab_carton_0054 | 6/5/24 | 6/22/24 | | H5-specific | TX |
| dholab_carton_0055 | 6/5/24 | 6/20/24 | | H5-specific | WI |
| dholab_carton_0056 | 6/5/24 | 6/18/24 | | H5-specific | WI |
| dholab_carton_0057 | 6/17/24 | 7/31/24 | | H5-specific | MN |
| dholab_carton_0058 | 6/17/24 | 7/8/24 | | H5-specific | IN |
| dholab_carton_0059 | 6/17/24 | 6/25/24 | | H5-specific | WI |
| dholab_carton_0060 | 6/17/24 | 7/3/24 | | H5-specific | WI |
| dholab_carton_0061 | 6/17/24 | 6/18/24 | | H5-specific | WI |
| dholab_carton_0062 | 6/17/24 | 7/3/24 | | H5-specific | WI |
| dholab_carton_0063 | 6/17/24 | 8/29/24 | 33 | H5-specific | MI |
| dholab_carton_0064 | 6/17/24 | 7/15/24 | | H5-specific | IL |
| dholab_carton_0065 | 6/17/24 | 8/23/24 | 29.89 | H5-specific | CO |
| dholab_carton_0066 | 6/17/24 | 7/1/24 | | H5-specific | IA |

| | | | | | |
|---|---|---|---|---|---|
| dholab_carton_0067 | 6/17/24 | 7/22/24 | | H5-specific | KY |

**S4 Table: SRA Accession numbers for sequences produced in this study.**

| Sample Name | SRA Accession |
|---|---|
| Carton 05 | SRR31094760 |
| Carton 06 | SRR31094759 |
| Carton 21 | SRR31094758 |
| Carton 24 | SRR31094757 |
| Carton 33 | SRR31094756 |
| Carton 36 | SRR31094755 |
| Carton 46 | SRR31094754 |
| Carton 48 | SRR31094753 |
| Carton 63 | SRR31094752 |
| Carton 65 | SRR31094751 |

**S5 Table: Acknowledgement to sequence contributors in Nextclade.**

| Author | sequences |
|---|---|
| Nguyen,T., Hutter,C., Markin,A., Thomas,M., Lantz,K., Killian,M.L., Janzen,G.M., Vijendran,S., Wagle,S., Inderski,B., Magstadt,D.R., Li,G., Diel,D.G., Frye,E.A., Dimitrov,K.M., Swinford,A.K., Thompson,A.C., Snevik,K.R., Suarez,D.L., Spackman,E., Lakin,S.M., C,S., Johnson,K.R., Baker,A.L., Robbe-Austerman,S., Torchetti,M.K., Anderson,T.K., Anderson,T. | 228 |
| Aufderhar,M., Franzen,K., Killian,M.L., Lantz,K., Stuber,T., Hicks,J., Norris,C., Lakin,S. | 261 |
| Kirby,M., Sheffield,S., Liddell,J., Frederick,J.C., Di,H., Lacek,K., Presley,S.M., Reinoso Webb,C., Malaeb,S., Cofi,M., Davis,T., Garten,R., Steel,J., Pusch,E., Barnes,J., Dugan,V., Garten,R.R. | 1 |
| Aufderhar,M., Franzen,K., Killian,M., Lantz,K., Stuber,T., Hicks,J., Norris,C., Lakin,S. | 575 |
| ? | 388 |
| Guan,L., Eisfeld,A.J., Pattinson,D., Gu,C., Biswas,A., Maemura,T., Trifkovic,S., Babujee,L., Presler,R. Jr., Dahn,R., Halfmann,P.J., Barnhardt,T., Neumann,G., Thompson,A., Swinford,A.K., Dimitrov,K.M., Poulsen,K., Kawaoka,Y., Presler,R. | 9 |
| Hicks,J.A., Stuber,T., Norris,C., Lakin,S., Killian,M.L., Franzen,K., Aufderhar,M. | 31 |
| Norris,C., Tod,S., Jessica,H., Matthew,A., Kerrie,F., Mary,K., Steven,L., Kris,L. | 63 |
| Messer,K., Gnirke,A., McMahon,K., Gomez,M., Stetson,L., Stachler,E., Hill,T., Hill,S., Knoll,H., Curtis,E., Samani,P., Wewior,N., Vuyk,W., O'Connor,D., Brown,C., Madoff,L., Wohl,S., Ozonoff,A., Park,D., MacInnis,B., Sabeti,P. | 19 |
| Halwe,N.J., Cool,K., Breithaupt,A., Schon,J., Trujillo,J.D., Nooruzzaman,M., Kwon,T., Ahrens,A.K., Britzke,T., McDowell,C.D., Piesche,R., Singh,G., Pinho dos Reis,V., Kafle,S., Pohlmann,A., Gaudreault,N., Corleis,B., Matias Ferreyra,F., Carossino,M., Balasuriya,U.B.R., Hensley,L., Morozov,I., Covaleda,L.M., Diel,D., Ulrich,L., Hoffmann,D., Beer,M., Richt,J.A. | 1 |
| Halwe,N.J., Ulrich,L., Schoen,J., Piesche,R., Britzke,T., Ahrens,A.K., Pinho dos Reis,V., Grund,C., Pohlmann,A., Breithaupt,A., Hoffmann,D., Beer,M., Harder,T. | 1 |
| Fabrizio,T.P., Kieffer,J.D., Franks,J., Jeevan,T., Kandeil,A., Walker,D., DeBeauchamp,J., Lowe,J.F., Bowman,A.S., Webby,R.J. | 17 |
| Barnes,J., Steel,J., Garten,R., Davis,T., Dugan,V., Riner,D.A., Soehnlen,M., Dover,M., Kirby,M.K., Sheffield,S., Liddell,J., Frederick,J.C., Di,H., Pusch,E., De La Cruz,J., Lacek,K.A. | 1 |
| Fabrizio,T.P., Kieffer,J.D., Franks,J., Jeevan,T., DeBeauchamp,J., Lowe,J.F., Bowman,A.S., Webby,R.J. | 3 |
| Roychoudhury,P., Han,P., Kong,K., Xie,H., Gamboa,L., Rodriguez-Salas,L., Ellis,S.E., Greninger,A., Bedford,T., Starita,L., Chu,H. | 1 |

| | |
|---|---|
| Suarez,D.L., Goraichuk,I.V., Killmaster,L., Spackman,E., Clausen,N.J., Colonius,T.J., Leonard,C.L., Metz,M.L. | 1 |
| Nguyen,T., C,C., Markin,A., Thomas,M., Lantz,K., Killian,M.L., Janzen,G.M., Vijendran,S., Wagle,S., Inderski,B., Magstadt,D.R., Li,G., Diel,D.G., Frye,E.A., Dimitrov,K.M., Swinford,A.K., Thompson,A.C., Snevik,K.R., Suarez,D.L., Spackman,E., Lakin,S.M., C,S., Johnson,K.R., Baker,A.L., Robbe-Austerman,S., Torchetti,M.K., Anderson,T.K., Anderson,T. | 1 |
| Burrough,E.R., Magstadt,D.R., Petersen,B., Timmermans,S.J., Gauger,P.C., Zhang,J., Siepker,C., Mainenti,M., Li,G., Thompson,A.C., Gorden,P.J., Plummer,P.J., Main,R., Hu,X. | 4 |
| Kirby,M.K., Steel,J., Garten,R., Davis,T., Dugan,V., Wilson,M., Liddell,J., Frederick,J.C., Di,H., Radford,K., De La Cruz,J., Keong,L., Lacek,K.A., Shu,B., Lopez,D., Seliskar,L., Le,H., Cadiz,V., Hua,C., Hok,S., Millena,S. | 1 |
| Kirby,M.K., Steel,J., Garten,R., Davis,T., Dugan,V., DaSilva,J., Liddell,J., Frederick,J.C., Di,H., Sheffield,S., Radford,K., De La Cruz,J., Keong,L., Lacek,K.A., Shu,B., Lopez,D., Seliskar,L., Le,H., Cadiz,V., Hua,C., Hok,S., Millena,S. | 4 |
| Kirby,M.K., Steel,J., Garten,R., Davis,T., Dugan,V., Sheffield,S., Liddell,J., Frederick,J.C., Di,H., Radford,K., De La Cruz,J., Keong,L., Lacek,K.A., Shu,B., Lopez,D., Seliskar,L., Le,H., Cadiz,V., Hua,C., Hok,S., Millena,S. | 1 |
| Oguzie,J.U., Marushchak,L.V., Shittu,I., Lednicky,J.A., Miller,A.L., Hao,H., Nelson,M.I., Gray,G.C., Rodriguez,A. | 1 |
| Shittu,I., Silva,D., Oguzie,J.U., Marushchak,L.V., Olinger,G., Lednicky,J.A., Trujillo-Vargas,C.M., Schneider,N.E., Hao,H., Gray,G.C. | 4 |
| Singh,G., Trujillo,J.D., Mcdowell,C., Fitz,I., Noll,L., Gaudreault,N., Retallick,J., Richt,J. | 1 |
| Barnes,J., Steel,J., Garten,R., Davis,T., Dugan,V., Matzinger,S.R., Kirby,M.K., Sheffield,S., Liddell,J., Frederick,J.C., Di,H., Pusch,E., De La Cruz,J., Keong,L., Lacek,K.A. | 4 |
